# Estimating Excess Mortality Among People Living with HIV/AIDS During the COVID-19 Pandemic in the USA

**DOI:** 10.1101/2025.06.09.25329225

**Authors:** Lindsay M. Hall, Hamed Karami, Gerardo Chowell

**Affiliations:** Department of Population Health Sciences, Georgia State University, Atlanta, GA, USA; Department of Mathematics and Statistics, Georgia State University, Atlanta, GA, USA; Department of Applied Mathematics, Kyung Hee University, Yongin, 17104, Korea

**Keywords:** HIV/AIDS, excess mortality, COVID-19, forecasting, health disparities, syndemic, United States

## Abstract

**Background:** Understanding syndemic effects of COVID-19 and HIV/AIDS can inform future public health preparedness. This study quantifies the all-cause excess death rate of people living with HIV/AIDS (PWHA) during the 2020-2022 COVID-19 pandemic in the United States (U.S.), including stratifications by sex, age, race/ethnicity, and region.

**Methods:** Using publicly available data from the CDC AtlasPlus dashboard, we employed ensemble n-subepidemic modeling framework (*SubEpiPredict* toolbox). This dynamic parametric approach is well suited for counterfactual forecasting with relatively short time series and was used to generate expected deaths among PWHA for 2020–2022 based on pre-pandemic trends. The models were calibrated using 12 years of pre-pandemic mortality trends (2008-2019), with the median excess death rate calculated as the difference between forecasted and observed death rates.

**Results:** Overall excess mortality among PWHA was estimated at 7,783 crude excess deaths (95% prediction interval [PI]:5,098–10,525), corresponding to 2.77 excess deaths per 100,000 people (95% PI:1.81–3.75), with the largest burden observed in 2021. Excess death rates were highest among male (3.39), age 55–64 (4.94), multiracial (12.82), and Northeast (4.12) groups. The largest crude number of excess deaths occurred among male (4,692), age 65 + (2,560), Black/African American (3,969), and Southern (4,025) groups.

**Conclusions:** These systematic, model-based results reveal stark heterogeneities among PWHA by exposing recent mortality patterns that may not be captured by disease-specific mortality reporting alone. These findings provide a robust, reproducible benchmark for assessing pandemic-related disparities and can inform future public health resource allocation and tailored interventions for vulnerable populations.

**Summary:** A study of people living with HIV/AIDS in the United States found substantial excess mortality during COVID-19, with marked disparities by age, race, sex, and region.

## Introduction

The convergence of two public health crises—COVID-19 and HIV— raises important questions about the intersectional impact of COVID-19 on people living with HIV/AIDS (PWHA), especially within already marginalized groups. Between 2020 and 2022, COVID-19 was listed as an underlying or contributing cause of over one million U.S. deaths—approximately 10.8% of all-cause mortality during that period [1]. Meanwhile, the U.S. mortality rate due to HIV has continued its gradual decline, dropping from 2.6 per 100,000 in 2010 to 1.3 per 100,000 in 2021 [2]. Despite this progress, HIV-related deaths remain disproportionately concentrated among specific populations, particularly males (76%), Black/African American individuals (49%), residents of the South (55%), and adults aged 45 or older (73%) [2]. Examining excess mortality among PWHA during the pandemic—across demographic groups—is essential to understanding potential compounded disparities and to inform more equitable public health responses in the future [3,4].

Evidence for increased mortality risk among PWHA stems from both biological vulnerabilities—such as potential immunosuppression due to low CD4 cell counts [5,6]—and a range of indirect pandemic effects, including reduced access to care and service interruptions [6,7]. Several studies report a moderately increased risk of COVID-19 mortality among people living with HIV (PWH) (Hazard ratios of 1.60-2.08) [5,8,9].

Overwhelmed healthcare systems diverted resources away from routine HIV prevention, testing, and treatment, while lockdowns and clinic closures disrupted antiretroviral therapy (ART) adherence and viral suppression efforts—especially in already resource-constrained communities [4,7]. These indirect effects likely contributed to elevated HIV/AIDS mortality during this period—deaths often termed “second-order deaths”—that may not be fully captured in standard cause-of-death records [7,10]. Viewed through a syndemic lens, the interaction between HIV and COVID-19 extends beyond biological co-infection risks to encompass compounding structural and social stressors, including reduced access to healthcare, social disruption, and economic instability [11].

Standard cause-of-death records are often insufficient to capture this complexity, as misclassification and underreporting are common during public health emergencies [12,10]. In this context, all-cause excess mortality serves as a more comprehensive metric, capable of capturing both direct and indirect deaths, including those not explicitly attributed to HIV/AIDS or COVID-19 [13].

Prior studies estimating excess deaths have relied on simple mathematical calculations among people living with HIV (PWH) [6,7]. However, they exclude regional stratifications and do not include deaths coded among people living with AIDS (PWA). Importantly, these analyses are restricted to the year 2020, capturing only the initial phase of the pandemic and omitting the broader compounding effects of subsequent waves, healthcare disruptions, and policy shifts [14,6,7].

Understanding demographic patterns in excess deaths is critical for identifying disparities and improving public health preparedness. This study addresses gaps in prior estimates by applying n-sub-epidemic modeling to quantify excess mortality among PWHA in the U.S. during the COVID-19 pandemic (2020-2022), stratified by sex, age, race/ethnicity, and region. By generating model-based counterfactual forecasts from pre-pandemic trends, this approach captures both direct and indirect mortality impacts while accommodating nonlinear dynamics in relatively short time series, while also contributing to the growing validation of the modeling framework itself [3,15,16,17]. These estimates provide a systematic and reproducible benchmark for assessing pandemic-related disparities and extend prior work by offering multi-year, demographically stratified insights to inform future public health response strategies.

## Materials and Methods

### Data Collection and Preparation

Yearly all-cause death rates and crude deaths (2008–2022) for PWHA were obtained from the CDC NCHHSTP AtlasPlus dashboard (accessed on February 25^th^, 2025), which provides overall and one-way stratified mortality data (i.e., stratified by a single demographic variable at a time). This database compiles de-identified HIV/AIDS surveillance data reported to the CDC through the National HIV Surveillance System (NHSS) by demographic category. CDC provides additional data collection details in their *Technical Notes [18].* At the time of analysis, death data for 2022 were considered preliminary and based on death data received by the CDC as of December 2023. Since the data is publicly accessible and de-identified, Institutional review board approval was not required.

### Excess Mortality Estimation

To estimate all-cause excess mortality among PWHA, we compared observed deaths during years 2020–2022 to counterfactual forecasts generated using the ensemble n-sub-epidemic modeling framework [15] calibrated to the pre-pandemic period (2008–2019). We computed both excess death rates and crude excess deaths yearly by demographic category.

Because AtlasPlus data are available only as overall or one-way stratified summaries, models were fitted separately for each stratum rather than jointly across demographic dimensions; consequently, stratified estimates are internally consistent but not additive across strata. The original dataset distinguished between all-cause deaths of PWH versus PWA; this would be expected to have relevance on mortality rates. Thus, for each demographic category, three separate (3) models were generated across disease indicators: 1) deaths among PWH, 2) deaths among PWA, and 3) combined deaths among PLHA.

The dynamic of the n-sub-epidemic framework is described by the following ODE system:

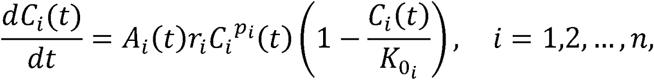

In this model, *C_i_* represents cumulative number of deaths, *r_i_* denotes growth rate, *p_i_* is scale of growth and 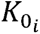 is epidemic size for the *i_th_* sub-epidemic, where 1≤ *i* ≤ *n*. Given *A*_1_(*t*) = 1, the indicator function *A_i_* characterizes the onset timing of the rest, *i_th_* sub-epidemic and is defined by:

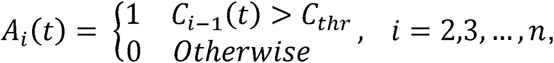

Where threshold *C_thr_* triggers sub-epidemic *i* when *C*_*i*-1_(*t*) > *C_thr_*. Therefore, the initial condition in this model is given by (*C*_1_(0),0,…,0) where *C*_1_(0) is the first reported data. The threshold values *C_thr_* are obtained by evenly dividing the cumulative sum of the smoothed signal into the same number of levels as there are data points. This method treats *C_thr_* as a series of possible threshold values, allowing the model to be tested at each fixed level instead of estimating the threshold directly from the data. The corrected Akaike Information Criterion (*AIC_c_*) is then used to identify the best-performing sub-epidemic models. Specifically, *AIC_c_* values are computed for the best-fitting models corresponding to different *C_thr_* values. *AIC_c_* is calculated as follows [19]:

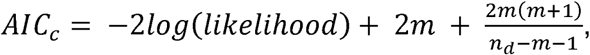

where *m* is the total number of data points and *n_d_* is the total number of data points calibrated.

Although annual HIV-related death rates exhibit relatively slowly changing trends, the n-sub-epidemic framework remains appropriate because its primary purpose is to generate probabilistic (uncertainty-quantified) forecasts, rather than to identify distinct epidemic waves. The n-sub-epidemic framework captures nonlinear trends, offers semi-mechanistic interpretability, and supports uncertainty quantification, making it well suited for counterfactual forecasting with limited data. Even under approximately linear baselines, the framework provides principled probabilistic estimates against which deviations during 2020–2022 can be formally assessed, with model flexibility constrained (maximum two sub-epidemics) to avoid overfitting. Across strata, the best-fitting models included both one- and two-sub-epidemic formulations.

### Model Calibration and Forecasting

Models were calibrated using observed death rates from 2008–2019 (baseline period). Model fitting was conducted using nonlinear least squares, assuming normally distributed errors. We assessed distributional assumptions using residual diagnostics (normal Q–Q plots and residuals versus fitted values). Residuals approximated a normal distribution, with minor departures at the extreme tails consistent with empirical aggregated mortality data. Of note, the model demonstrated good agreement between fitted and observed pre-pandemic trends, supporting its suitability for generating counterfactual forecasts.

Yearly forecasts of death rates by demographic category were then generated for 2020-2022. Models include a median trendline of death rate, as well as a 95% prediction interval (PI) to capture uncertainty. The 95% prediction interval bounds of the median death rates were used to compute the excess death rate intervals, using previously established methodology [16,20], as described below.

Let ŷ⍰represent the forecasted death rate per 100,000 population in year t ∈ {2020, 2021, 2022}, and y⍰ the corresponding observed death rate. The annual excess death rate (EDR) is calculated as:

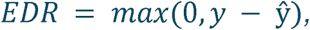

where EDR⍰ denotes the excess death rate per 100,000 people for year t compared to expected levels based on pre-pandemic trends. The non-negativity constraint reflects the standard interpretation of excess mortality as deaths above an expected counterfactual baseline. However, for cumulative burden estimation, negative values were set to zero so that periods of lower-than-expected mortality do not offset excess deaths. Here, “burden” refers to total deaths above the expected baseline (not net change), consistent with standard excess mortality reporting [20]. Thus, results should be interpreted as estimates of excess mortality burden rather than net mortality deviation. Since our forecasted trend ŷ⍰includes a 95% prediction interval (PI), the lower and upper bounds of the excess death rate are computed as follows:

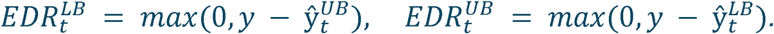

Hence, we ensure that the uncertainty from the forecast is propagated into the excess mortality estimates.

To calculate crude excess deaths, each excess death rate was multiplied by the respective population subgroup size NLZ given in the original dataset:

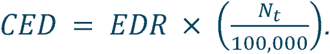

Lower and upper bounds for crude excess deaths were calculated analogously, using the corresponding bounds of EDRLZ.

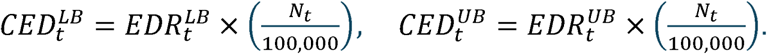

It is worth noting that this method allows for quantifying excess mortality across demographic groups while incorporating forecast uncertainty. Importantly, excess death rates are interpreted as the additional deaths per 100,000 people beyond expected mortality, capturing both direct and indirect effects of the pandemic.

### Ensemble Construction

A weighted (W) ensemble was derived from the n-sub-epidemic framework to produce all forecasts [15]. This framework aggregates multiple asynchronous sub-epidemic models and creates forecasted trajectories. In this analysis, sub-epidemics are modeled as generalized logistic growth models and assume a maximum of 2 sub-epidemics in the trajectory to reduce overfitting and improve stability, given the limited number of annual observations. Model parameters are estimated using the nonlinear least squares method. To estimate parameters with known uncertainty, 300 bootstrap samples per model were generated using the approach described by Hastie et al. [21].

We constructed two ensemble model variations from the top two ranked models based on *AIC_c_*. We restricted the ensemble to the top two ranked models to preserve parsimony and forecast stability given the short annual time series. The weighted (W) ensemble model utilized relative likelihood to determine the weights of the top two ranked models, and the unweighted (UW) ensemble model assumed equal weight for the top two ranked models.

### Software

Analyses were performed using MATLAB (R2023a).

## Results

Primary results presented in this study are derived from the weighted (W)ensemble models due to their superior calibration performance. Trends from unweighted (UW) models closely mirrored those of W models but exhibited wider uncertainty intervals; full UW results are provided in Supplementary Figures S1–S5.

### Overall Patterns

Weighted (W) ensemble models forecast a continued decline in death rates for HIV, AIDS, and combined HIV/AIDS during the pandemic period (Figure 1, Supplementary Figures S6-S9). In contrast, observed death rates during 2020–2022 exceeded the model-based 95% prediction intervals (PI) across all disease categories. Excess mortality peaked in 2021, with estimated increases of 1.80 (95% PI: 1.23–2.39),1.19 (95% PI: 0.80–1.61), and 2.77 (95% PI: 1.81–3.75) deaths per 100,000 population above expected pre-pandemic levels for respective HIV, AIDS, and combined HIV/AIDS (Figure 2). Correspondingly, crude excess deaths in 2021 were highest for the combined HIV/AIDS category at 7,793 deaths (95% PI: 5,098–10,525) (Supplementary Figure S10). Residual diagnostics supported the assumed error structure, with approximately normal residual distributions and minor departures at extreme values (Supplementary Figure S11).

**Figure 1.**
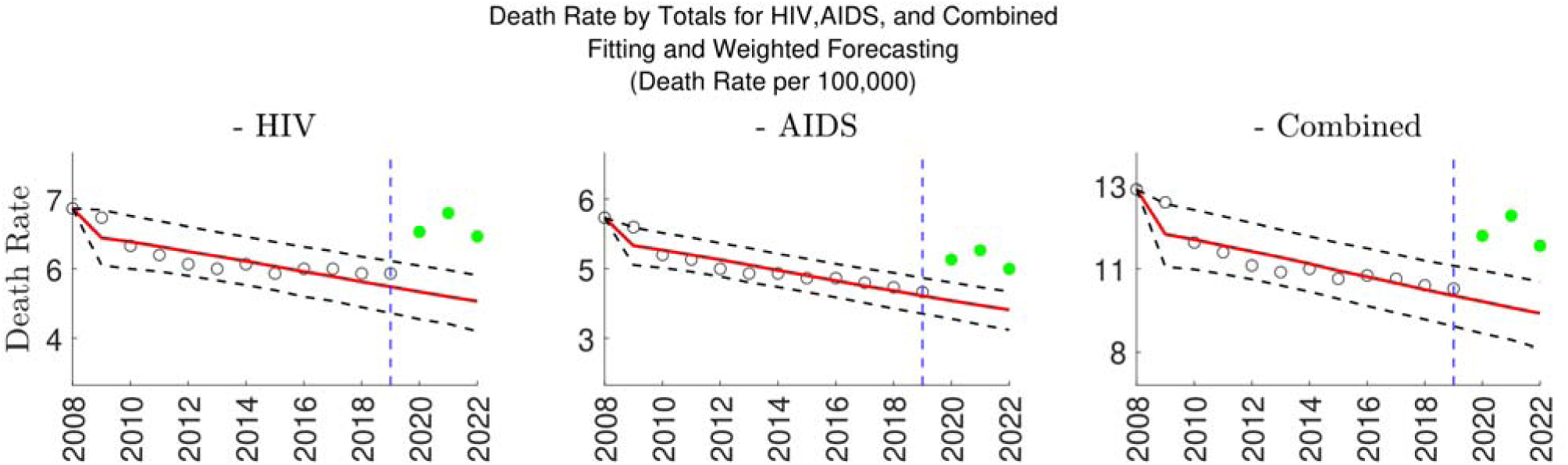
Death rates (deaths per 100,000 people) for HIV, AIDS, and HIV/AIDS combined by year using the weighted ensemble model. The solid red line represents the modeled median death rate, with the dotted black line representing 95% Prediction Interval (PI). Data points shown in black represent observed data used for the calibration period, while solid-filled green/yellow data points in years 2020-2022 represent forecasted data. Forecasted data in green represents excess death rates outside of the 95% PI, while forecasted data in yellow, if present, represents excess data rates within the 95% PI. The blue dotted vertical line denotes year 2019, which is the end of the calibration period, and before COVID-19.

**Figure 2.**
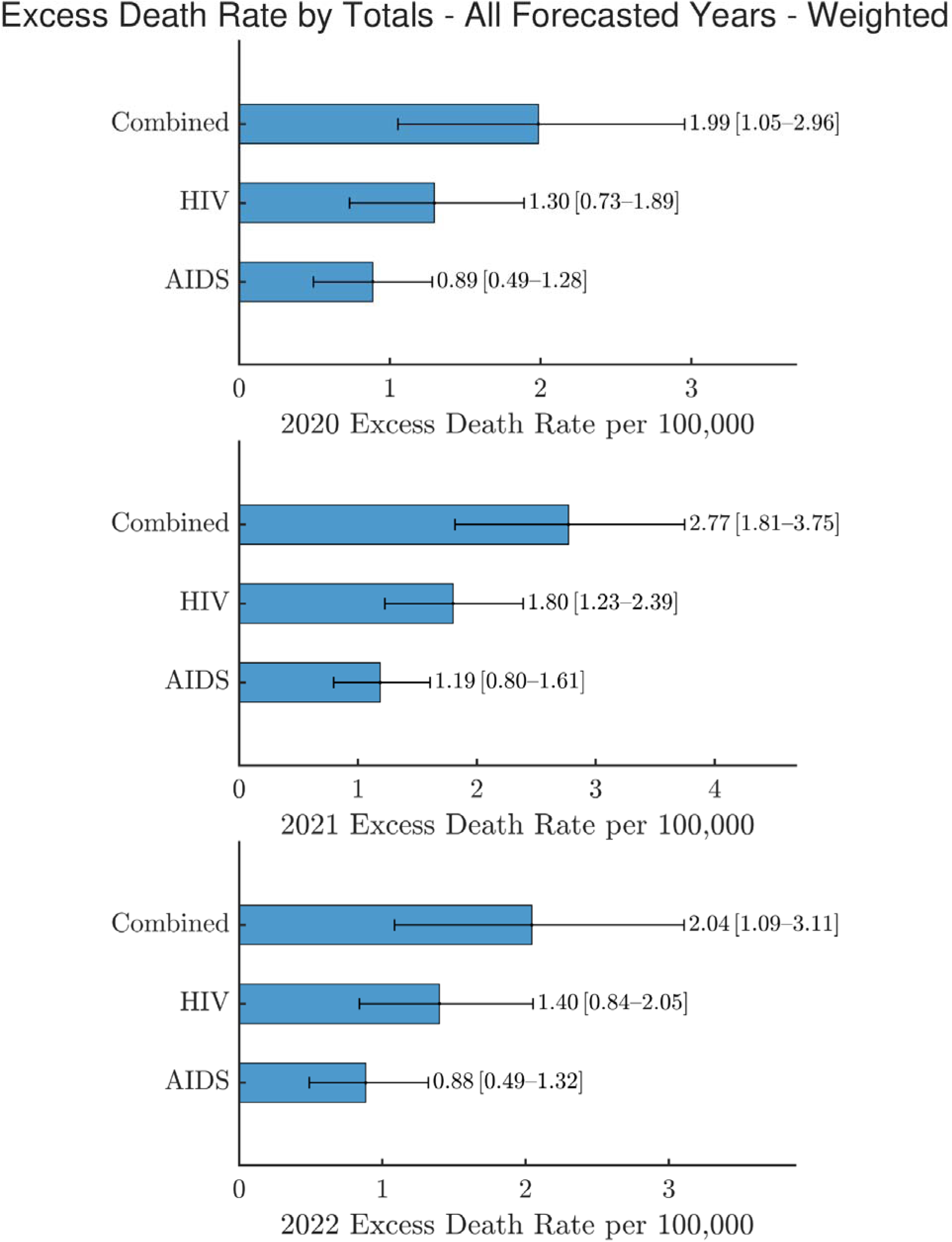
Excess death rate (per 100,000 people) for HIV, AIDS, and HIV/AIDS Combined for 2020-2022 Using Weighted Ensemble Forecasting. Top to bottom: years 2020, 2021, 2022. *The 95% upper and lower bounds for median excess death rates are displayed in black bars*.

### Stratified Results

By sex, males consistently experienced higher excess death rates than females across all disease indicators, with the largest excess observed in 2021 for the male combined HIV/AIDS group (3.39 per 100,000; 95% PI: 1.69–4.99) (Figure 3). Crude excess deaths among males in 2021 reached 4,692 (95% PI: 2,338–6,907) (Supplementary Figure S12).

**Figure 3.**
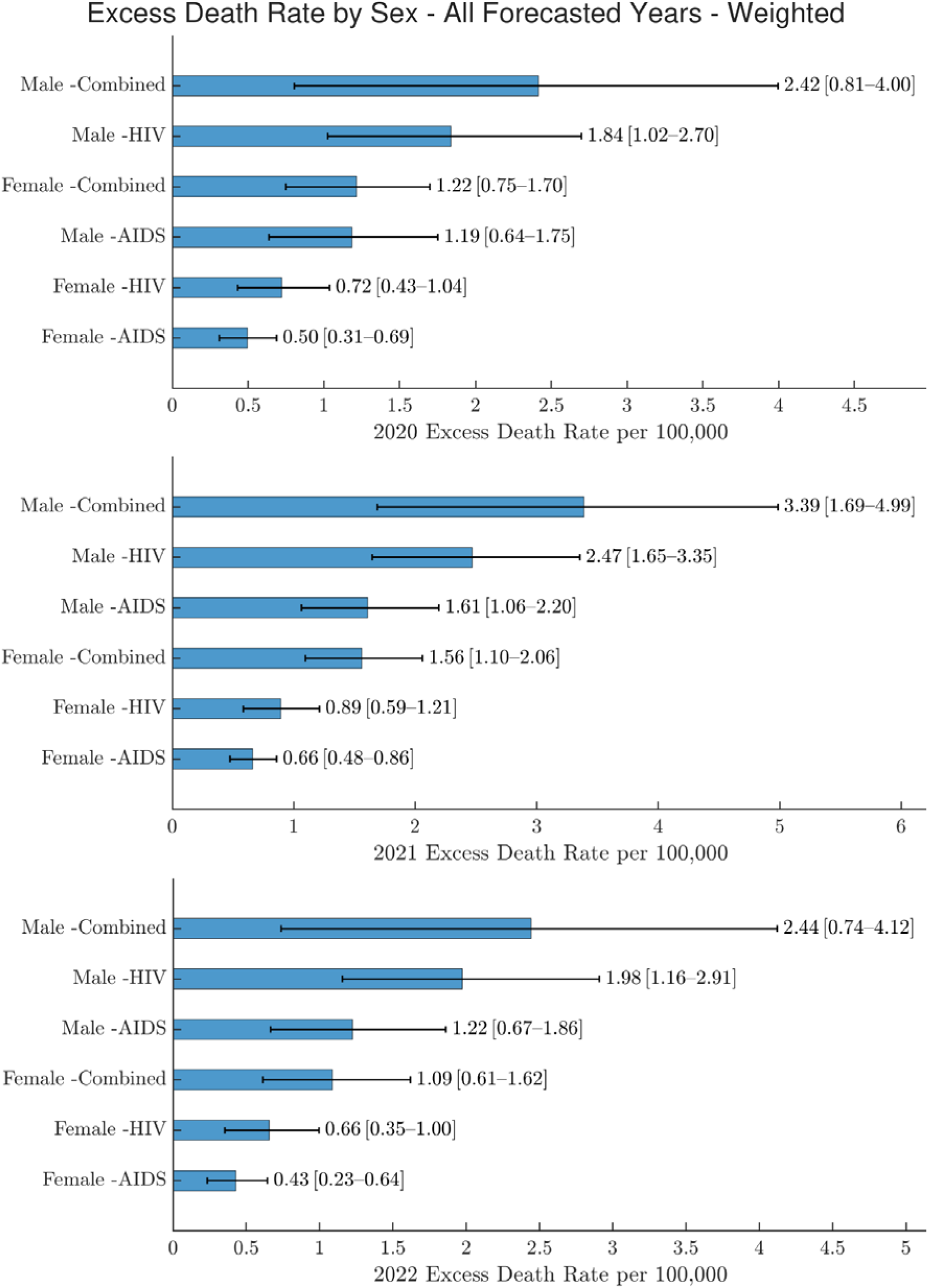
Excess death rate (per 100,000 people) for HIV, AIDS, and HIV/AIDS Combined by Sex for 2020-2022 Using Weighted Ensemble Forecasting. Top to bottom: years 2020, 2021, 2022. *The 95% upper and lower bounds for median excess death rates are displayed in black bars*.

Age-stratified analyses revealed the greatest excess mortality burden among older adults. Individuals aged 55–64 exhibited the highest excess death rate for combined HIV/AIDS (4.94 per 100,000; 95% PI: 3.24–6.68), while the largest absolute number of excess deaths occurred among those aged 65 years and older in 2021 (2,560 deaths; 95% PI: 1,391–3,616) (Figure 4; Supplementary Figure S13).In contrast, younger age groups exhibited minimal or zero excess mortality. Racial disparities were pronounced. Multiracial and Black/African American populations experienced the highest excess death rates for combined HIV/AIDS, at 12.82 (95% PI: 5.44–20.38) and 11.42 (95% PI: 7.07–16.10) per 100,000, respectively (Figure 5). The largest crude excess mortality burden occurred among Black/African American individuals in 2021, with 3,969 excess deaths (95% PI: 2,455–5,591) (Supplementary Figure S14).

**Figure 4.**
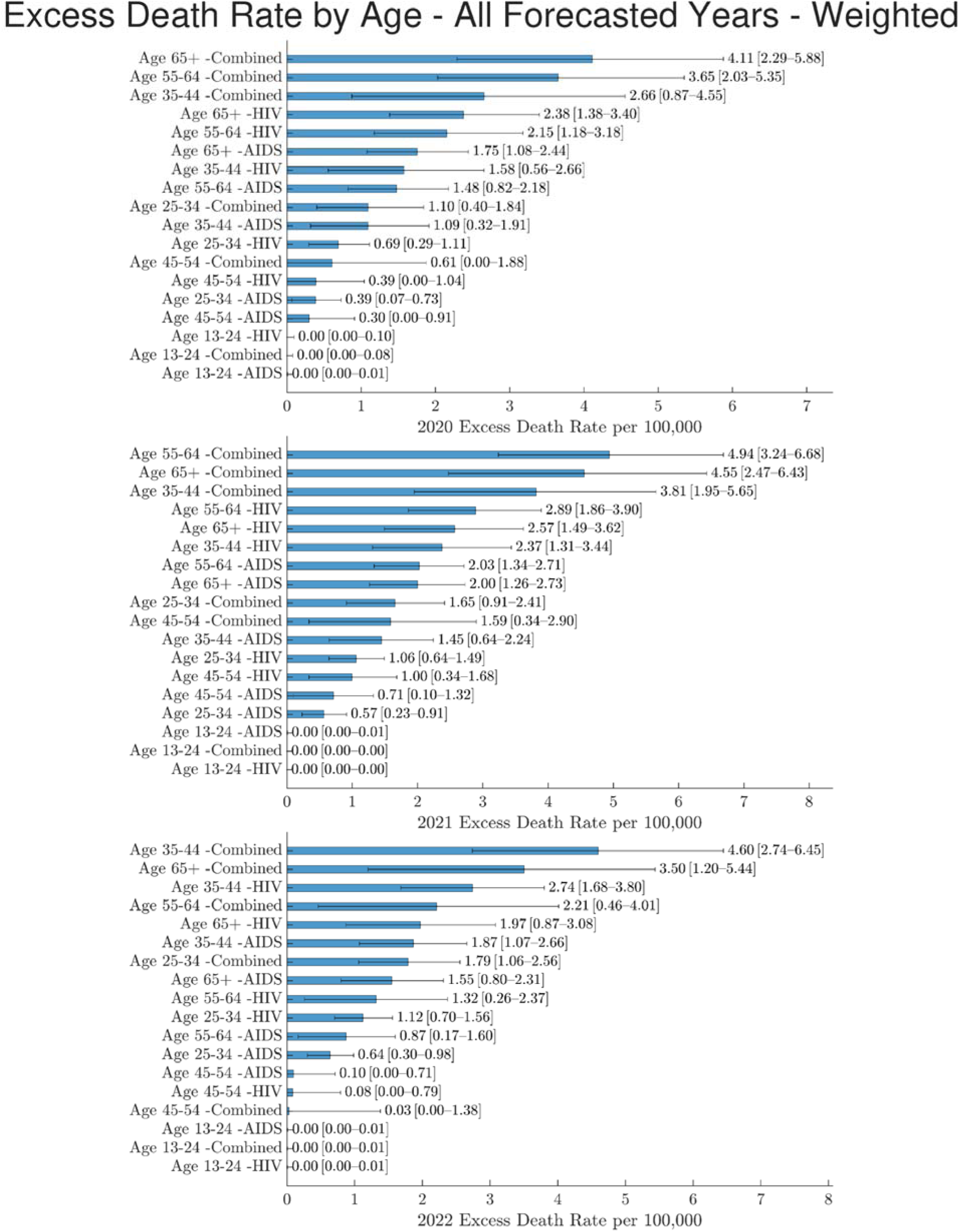
Excess death rate (per 100,000 people) for HIV, AIDS, and HIV/AIDS Combined by Age for 2020-2022 Using Weighted Ensemble Forecasting. Top to bottom: years 2020, 2021, 2022. *The 95% upper and lower bounds for median excess death rates are displayed in black bars*.

**Figure 5.**
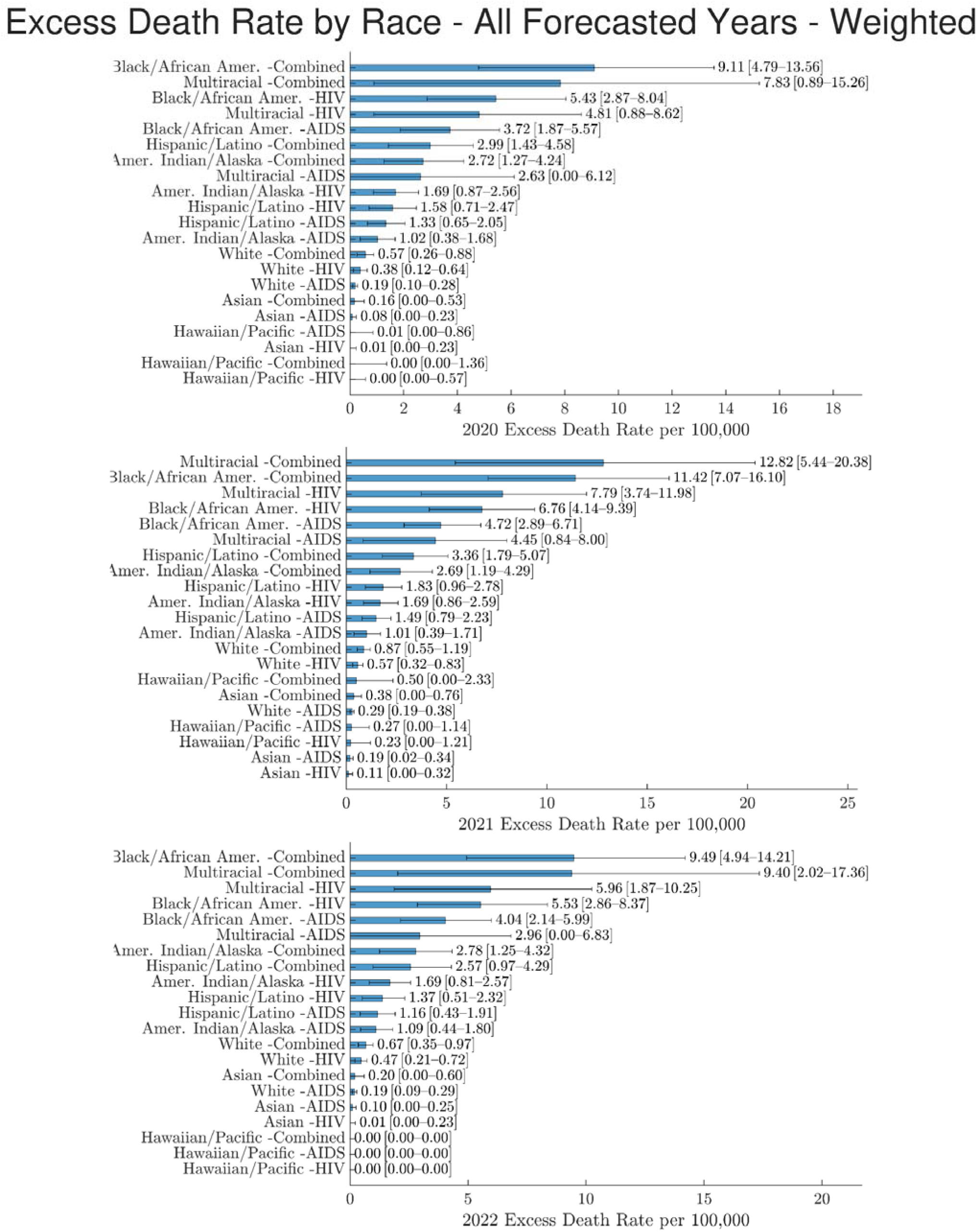
*Excess death rate (per 100,000 people) for HIV, AIDS, and HIV/AIDS Combined by Race for 2020-2022 Using Weighted Ensemble Forecasting.* Top to bottom: years 2020, 2021, 2022. *The 95% upper and lower bounds for median excess death rates are displayed in black bars*.

Regional analyses indicated heterogeneous timing and magnitude of excess mortality. The Northeast experienced the highest excess death rate in 2020 (4.12 per 100,000; 95% PI: 2.76–5.49), whereas the South accounted for the largest absolute number of excess deaths in 2021 (4,025 deaths; 95% PI: 2,450–5,729) (Supplementary Figures S15-S16).

## Discussion

Our results highlight stark heterogeneities in pandemic-related excess mortality. Overall, we estimated excess deaths among PWHA at 7783 crude excess deaths, or a rate of 2.77 excess deaths per 100,000 people, peaking in 2021. The highest excess death rates were observed among males (3.39), those aged 55-64 (4.94), multiracial (12.82), and among those in the Northeast U.S. region (4.12). When considering the U.S. population at large, crude deaths were highest among the following categories: Males (4,692), age group 65+ (2,560), Black/African Americans (3,969), and among those in the South U.S. region (4,025). Generally, amongst demographic groups in the forecasted pandemic period (2020-2022), excess death rates and crude estimates peak in year 2021, highlighting the effectiveness of ongoing dual interventions such as COVID-19 vaccine uptake and improved access to medical care for PWHA after COVID-19 pandemic disruptions settled in 2022 [22,23].

This analysis uniquely contributes to the literature by quantifying all-cause excess deaths among PWHA during the COVID-19 pandemic via an n-sub-epidemic forecasting method—stratified by race, sex, age, and region. The aforementioned syndemic conceptual framework [11] not only contextualizes the complex interplay of multiple epidemics but also enhances the interpretive value of our stratified excess mortality estimates. This analysis of all-cause excess mortality incorporates deaths that may have been exacerbated by other interacting syndemic risk factors and thus offers a nuanced quantitative view of how overlapping structural, social, and biological vulnerabilities may have compounded mortality risks in marginalized populations. Our stratified results underscore the need for tailored interventions in future pandemics, especially for populations already burdened by systemic health disparities.

### Demographic Disparities

Our study reinforces well-documented disparities in HIV-related outcomes across key demographic groups. HIV is well known to have a disproportionate impact on certain populations, particularly people of color, men who have sex with men, and transgender women. In 2022, Black Americans made up 40% of people living with HIV and 43% of HIV-related deaths, despite representing only 12% of the population. Similarly, Hispanic/Latino individuals comprised 26% of people living with HIV, exceeding their 19% share of the population [24]. The burden is especially severe in the Southern U.S., where eight of the ten states with the highest diagnosis rates are located—driven by limited Medicaid expansion, healthcare provider shortages, low health literacy, and HIV stigma [24,25]. These disparities persist in both infection rates and outcomes, with lower survival rates and higher HIV-related mortality among Black and Latino populations [24]. Well-established racial barriers to care have been identified as causes of these inequities, including disparate access, knowledge, and prioritization to PrEP and treatment as prevention (TasP) [26,27,28]. These disparities in HIV outcomes cannot be explained by access to care alone; they are also shaped by entrenched systemic issues like racism and stigma surrounding HIV [29,28,25].

While pre-existing demographic disparities embedded in baseline mortality are not inherently captured in excess death rate estimates, they may still contribute to higher observed death rates and thus amplify estimated excess mortality. In fact, these disparities may be amplified due to the dual burden of living with HIV/AIDS and experiencing the biological, psychological, social, and systemic impacts of COVID-19.

### Literature Comparisons

Comparisons to previously published estimated excess deaths can be made only for 2020, since prior research is limited to that year. When interpreting these comparisons, it is important to recognize key methodological differences. Previous estimates relied primarily on simple comparisons to short historical baselines and were restricted to people living with HIV, excluding individuals diagnosed with AIDS, whereas our approach generates probabilistic counterfactuals conditioned on long-term pre-pandemic trends and includes both PWH and PWA.

Compared with Viguerie et al. [7], our estimated all-cause crude excess deaths for 2020, when restricted to comparable demographic groups (i.e., individuals with HIV who are Black/African American, Hispanic/Latino, or White; all ages, sexes, and U.S. regions), was 3,296 (95% PI: 1,150–5,070) (Supplementary Figure S14), representing a 2.1-fold increase compared to Viguerie et al.’s estimate of 1,550 (95% CI: 1,229–1,877). This difference likely reflects differences in baseline construction, as Viguerie et al. compare 2020 outcomes to 2018–2019 mortality levels, whereas our framework forecasts counterfactual mortality using 12 years of pre-pandemic data (2008–2019) and explicitly propagates uncertainty. Across sex, age, and race strata, this ratio remains broadly consistent, with the upper bound of the Viguerie et al. CI generally falling near or within our 95% PI lower bound [7]. Only a limited comparison can be made to Zhu et al. [6], who estimate COVID-19–specific excess deaths among PWH, rather than all-cause excess mortality. Because their analysis includes all demographic groups without stratification, our 2020 crude excess death estimate (3,622, 95% PI: 2,050–5,287) reflects a 2.5-fold increase over their reported estimate of 1,457 (Supplementary Figure S10). This difference is expected, as our estimates capture both direct and indirect mortality effects, including deaths from non-COVID-19 causes exacerbated by pandemic-related healthcare disruptions.

Although Ashraf et al. [14] examined only standard (non-excess) HIV-coded death rates through 2020, we observe consistent qualitative trends across sex, age, race, and region when comparing their findings with the fitted and observed trajectories from our models, providing additional face validity.

Taken together, these comparisons indicate that our excess mortality estimates are higher than several previously published values, but this should not be interpreted as inconsistency or overestimation. Rather, our results represent upper-bound burden estimates that incorporate long-term pre-pandemic trends, multi-year pandemic effects, and uncertainty propagation, offering a complementary perspective to earlier, more conservative approaches. By applying a validated, model-based forecasting framework to all-cause mortality data, we extend prior efforts [15] by capturing trends over a longer time horizon and across key demographic groups, providing a transparent benchmark for future studies of pandemic-related disparities and syndemic impacts.

### Limitations

Our study is not exempt from limitations. Because AtlasPlus data are available only as overall or one-way stratified summaries, models were estimated independently by sex, age, race/ethnicity, and region; thus, stratified crude excess deaths are not expected to sum to the overall estimate and should be interpreted as parallel, internally consistent assessments. In addition, all-cause excess mortality could not be disaggregated by cause of death, precluding attribution to HIV/AIDS-, COVID-19-, or other cause-specific pathways [2].

Importantly, this study does not include a comparison group without HIV/AIDS; therefore, it does not estimate the causal effect of HIV status on excess mortality, but rather quantifies deviations from expected mortality trends within the PWHA population. However, excess mortality was widely observed across the general U.S. population during the COVID-19 pandemic, reflecting both direct and indirect impacts of the pandemic on mortality. Moreover, qualitatively similar patterns of excess mortality observed in other chronic disease populations during the COVID-19 pandemic suggest that healthcare disruptions broadly affected medically vulnerable groups [30]. Stratification by comorbidities (e.g., cardiovascular disease, diabetes, mental illness, or substance use disorders) was not feasible due to data aggregation, although these conditions are more prevalent among PWHA and likely contributed to excess mortality. Within this broader context, our findings highlight substantial heterogeneity in excess mortality among PWHA across demographic groups, rather than quantifying differences relative to the general population. Future work incorporating appropriate comparison groups would help clarify the extent to which HIV/AIDS contributed to excess mortality beyond broader pandemic-related effects.

Year-to-year comparisons, particularly between 2020 and 2021, should be interpreted cautiously, as pandemic-related disruptions to HIV testing, diagnosis, reporting, and care likely affected both observed deaths and population denominators, with higher excess mortality in 2021 reflecting delayed care, improved ascertainment, and accumulated indirect effects.

Additionally, this study was not designed to identify causal mechanisms underlying observed disparities in excess mortality across demographic groups. Although we discuss plausible explanations based on prior literature, the observed patterns should be interpreted cautiously, as they likely reflect a combination of pre-existing health disparities and pandemic-related disruptions in healthcare access, rather than direct causal effects that can be inferred from the present analysis.

### Conclusions

Our results underscore the urgent need to account for the compounded vulnerabilities faced by PWHA during public health crises. By applying a model-based excess mortality framework, we reveal recent mortality patterns that may not be captured by disease-specific mortality reporting alone, thereby quantifying the cumulative impact of overlapping epidemics. Understanding how structural inequities, healthcare disruptions, and broader biopsychosocial stressors influence mortality among PWHA can guide policymakers in designing more equitable and resilient emergency response plans. These findings provide actionable evidence to inform future public health programming, resource allocation, and policy decisions aimed at protecting PWHA and other medically vulnerable populations during future pandemics.

## Data Availability

All data produced in the present work are contained in the manuscript, via online access per included link.

https://github.com/lhall63/U.S.-Excess-Death-Rates-among-PLHA-during-COVID-19/

## Acknowledgements

1. Author Contributions: L.H. performed data analysis and contributed to manuscript drafting and editing. H.K. assisted with generating figures in MATLAB. G.C. conceived the project, provided general guidance, and revised the manuscript.
2. Research presented at Emory University Summer Institute in Statistics and Modeling in Infectious Diseases (SSMID) Conference. July 2025. Atlanta, Georgia, USA.
3. Original data & analytical code can be found at: https://github.com/lhall63/U.S.-Excess-Death-Rates-among-PLHA-during-COVID-19/

