## Supplementary Figures for "Estimating Excess Mortality Among People Living with HIV/AIDS During the COVID-19 Pandemic in the USA"

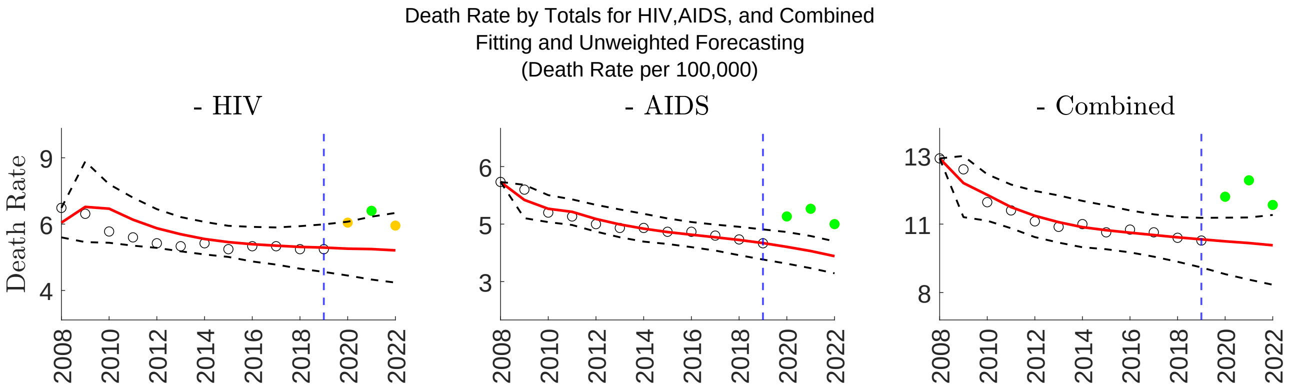


Figure S1: Death rate for HIV, AIDS, and HIV/AIDS combined by year using unweighted ensemble model. This figure represents overall death rate in deaths per 100,000 people. The solid red line represents the modeled median death rate, with the dotted black line representing 95% Prediction Interval (PI). Data points shown in black represent observed data used for the calibration period, while solid-filled green/yellow data points in the years 2020-2022 represent forecasted data. Forecasted data in green represents excess death rates outside of the 95% PI, while forecasted data in yellow, if present, represents excess data rates within the 95% PI. The blue dotted vertical line denotes the year 2019, which is the end of the calibration period, and before COVID-19.

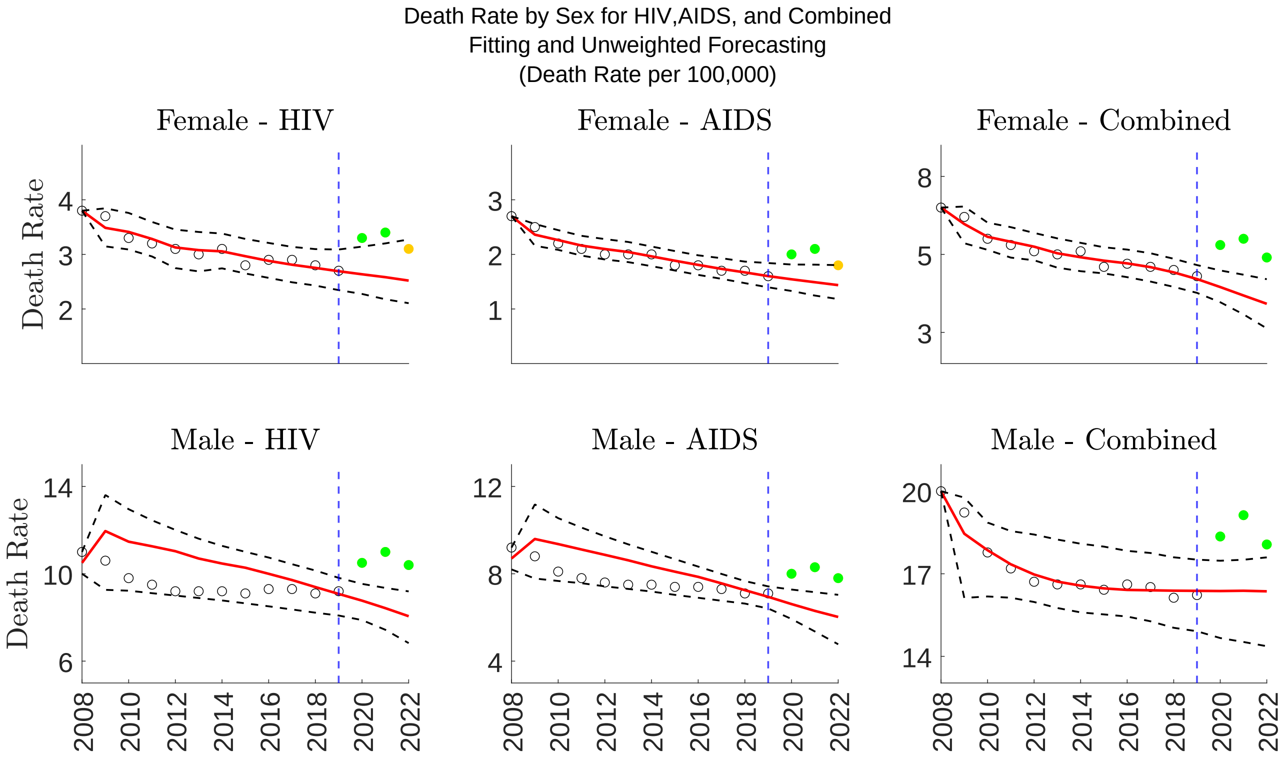


Figure S2: Death rate for HIV, AIDS, and HIV/AIDS combined by year using unweighted ensemble model, stratified by sex. This figure represents overall death rate in deaths per 100,000 people. The solid red line represents the modeled median death rate, with the dotted black line representing 95% Prediction Interval (PI). Data points shown in black represent observed data used for the calibration period, while solid-filled green/yellow data points in the years 2020-2022 represent forecasted data. Forecasted data in green represents excess death rates outside of the 95% PI, while forecasted data in yellow, if present, represents excess data rates within the 95% PI. The blue dotted vertical line denotes the year 2019, which is the end of the calibration period, and before COVID-19.

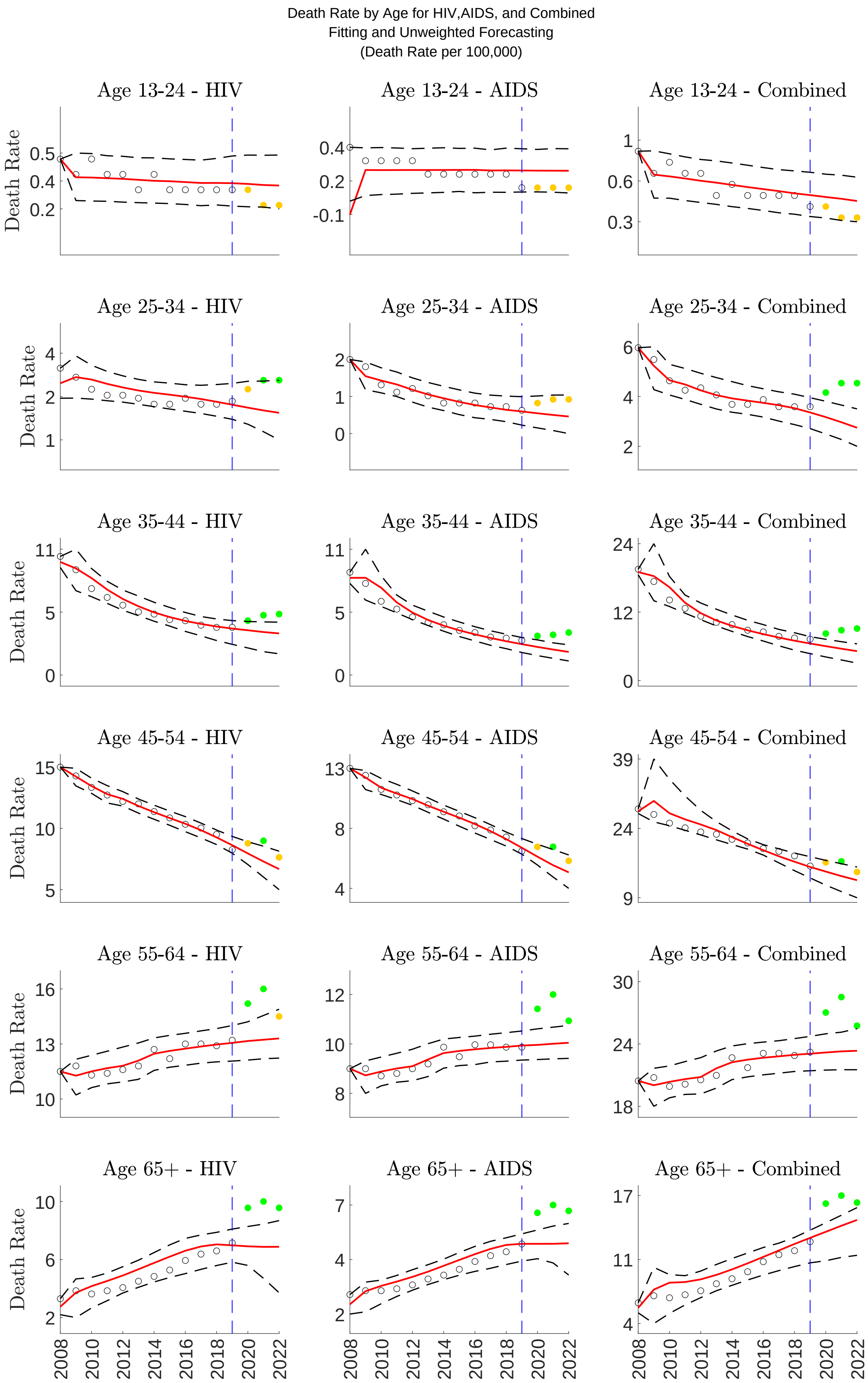


Figure S3: Death rate for HIV, AIDS, and HIV/AIDS combined by year using unweighted ensemble model, stratified by age. This figure represents overall death rate in deaths per 100,000 people. The solid red line represents the modeled median death rate, with the dotted black line representing 95% Prediction Interval (PI). Data points shown in black represent observed data used for the calibration period, while solid-filled green/yellow data points in the years 2020-2022 represent forecasted data. Forecasted data in green represents excess death rates outside of the 95% PI, while forecasted data in yellow, if present, represents excess data rates within the 95% PI. The blue dotted vertical line denotes the year 2019, which is the end of the calibration period, and before COVID-19.

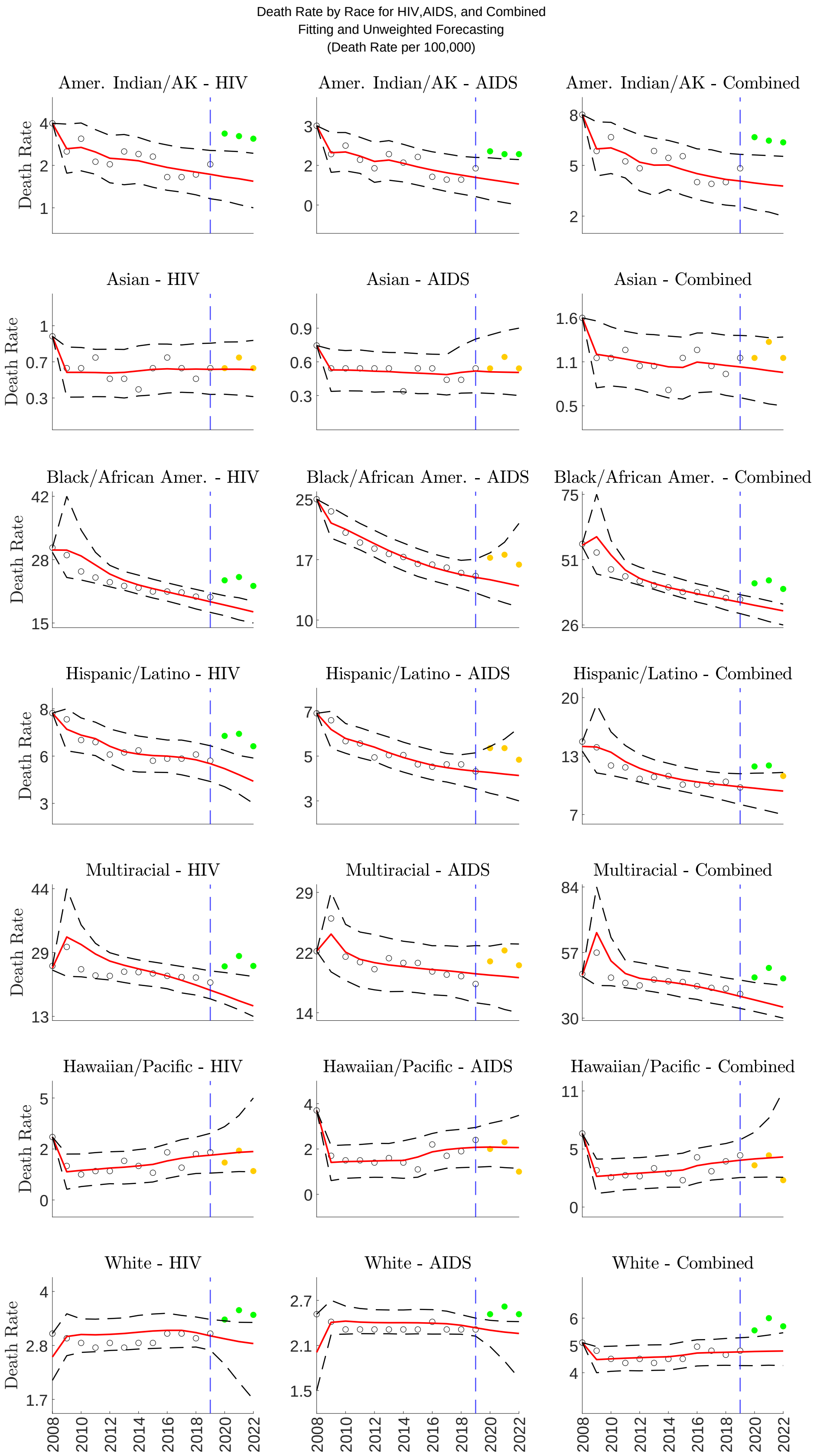


Figure S4: Death rate for HIV, AIDS, and HIV/AIDS combined by year using unweighted ensemble model, stratified by race. This figure represents overall death rate in deaths per 100,000 people. The solid red line represents the modeled median death rate, with the dotted black line representing 95% Prediction Interval (PI). Data points shown in black represent observed data used for the calibration period, while solid-filled green/yellow data points in the years 2020-2022 represent forecasted data. Forecasted data in green represents excess death rates outside of the 95% PI, while forecasted data in yellow, if present, represents excess data rates within the 95% PI. The blue dotted vertical line denotes the year 2019, which is the end of the calibration period, and before COVID-19.

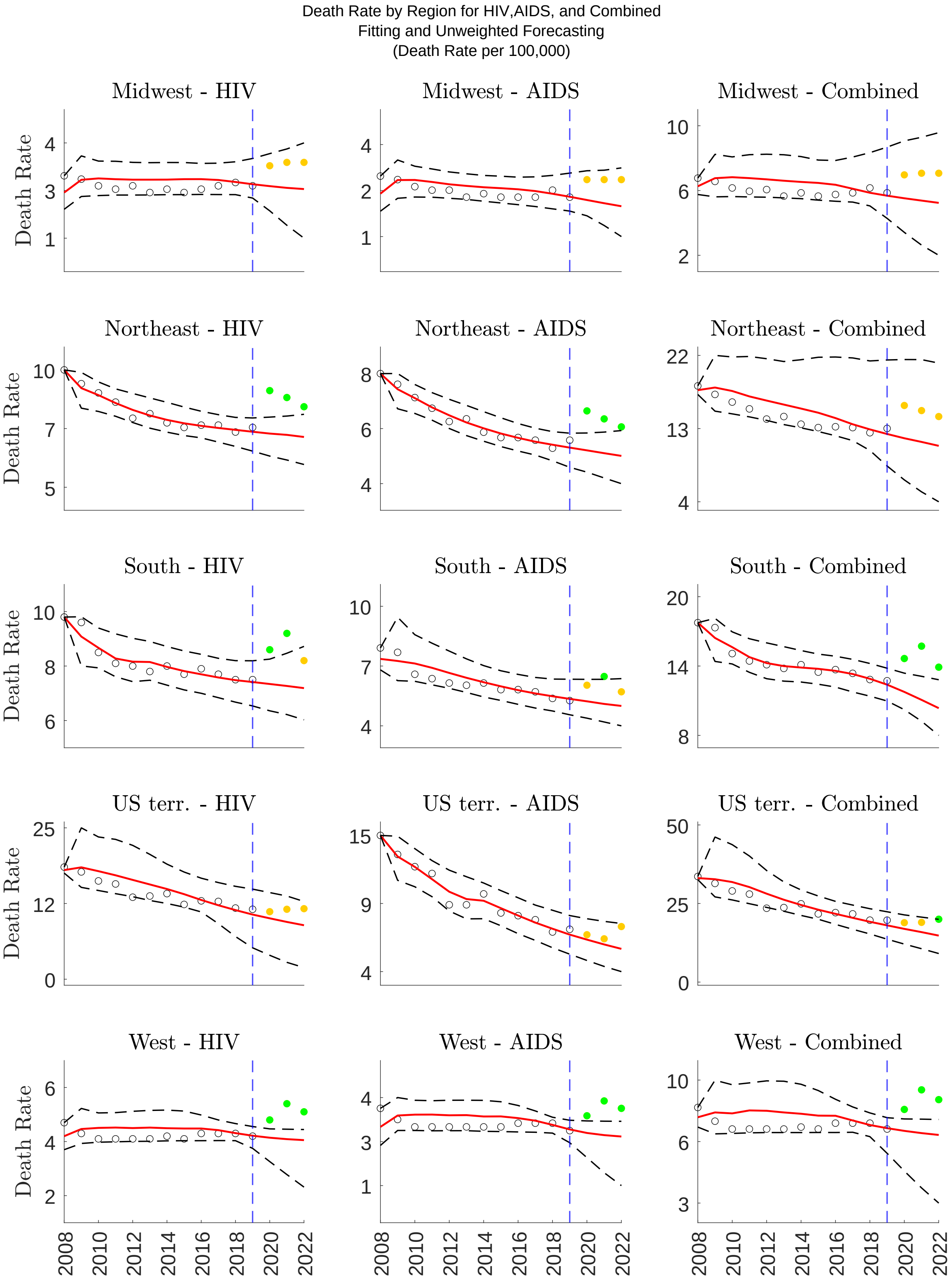


Figure S5: Death rate for HIV, AIDS, and HIV/AIDS combined by year using unweighted ensemble model, stratified by region. This figure represents overall death rate in deaths per 100,000 people. The solid red line represents the modeled median death rate, with the dotted black line representing 95% Prediction Interval (PI). Data points shown in black represent observed data used for the calibration period, while solid-filled green/yellow data points in the years 2020-2022 represent forecasted data. Forecasted data in green represents excess death rates outside of the 95% PI, while forecasted data in yellow, if present, represents excess data rates within the 95% PI. The blue dotted vertical line denotes the year 2019, which is the end of the calibration period, and before COVID-19.

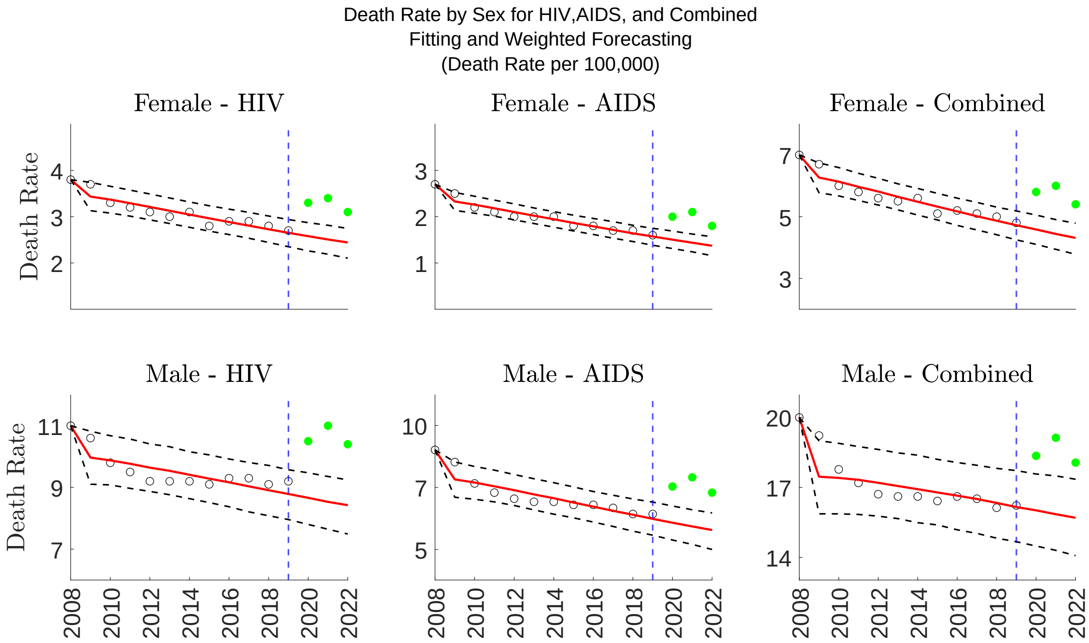


Figure S6: Death rate for HIV, AIDS, and HIV/AIDS combined by year using weighted ensemble model, stratified by sex. This figure represents overall death rate in deaths per 100,000 people. The solid red line represents the modeled median death rate, with the dotted black line representing 95% Prediction Interval (PI). Data points shown in black represent observed data used for the calibration period, while solid-filled green/yellow data points in the years 2020-2022 represent forecasted data. Forecasted data in green represents excess death rates outside of the 95% PI, while forecasted data in yellow, if present, represents excess data rates within the 95% PI. The blue dotted vertical line denotes the year 2019, which is the end of the calibration period, and before COVID-19.

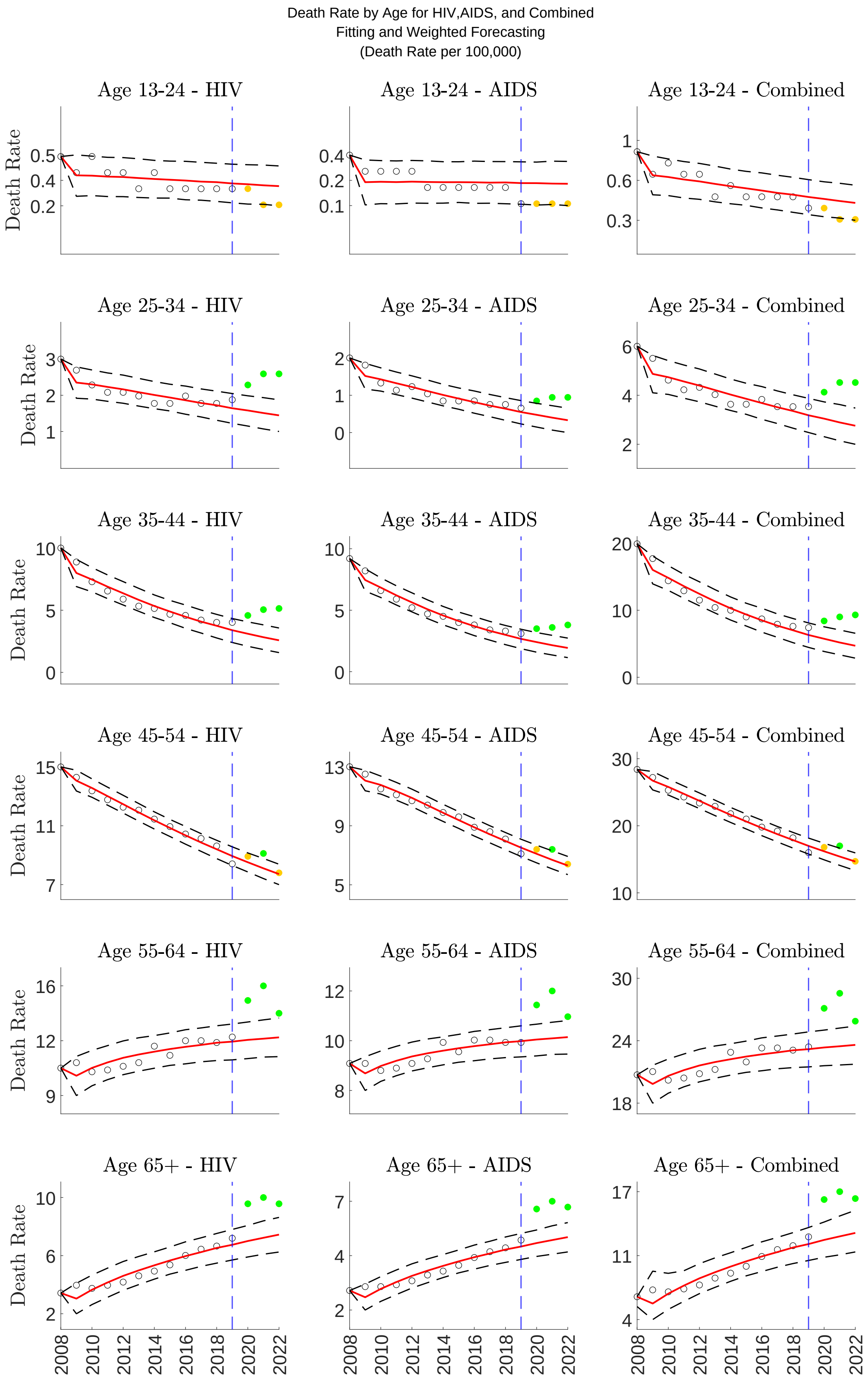


Figure S7: Death rate for HIV, AIDS, and HIV/AIDS combined by year Using weighted ensemble model, stratified by age. This figure represents overall death rate in deaths per 100,000 people. The solid red line represents the modeled median death rate, with the dotted black line representing 95% Prediction Interval (PI). Data points shown in black represent observed data used for the calibration period, while solid-filled green/yellow data points in the years 2020-2022 represent forecasted data. Forecasted data in green represents excess death rates outside of the 95% PI, while forecasted data in yellow, if present, represents excess data rates within the 95% PI. The blue dotted vertical line denotes the year 2019, which is the end of the calibration period, and before COVID-19.

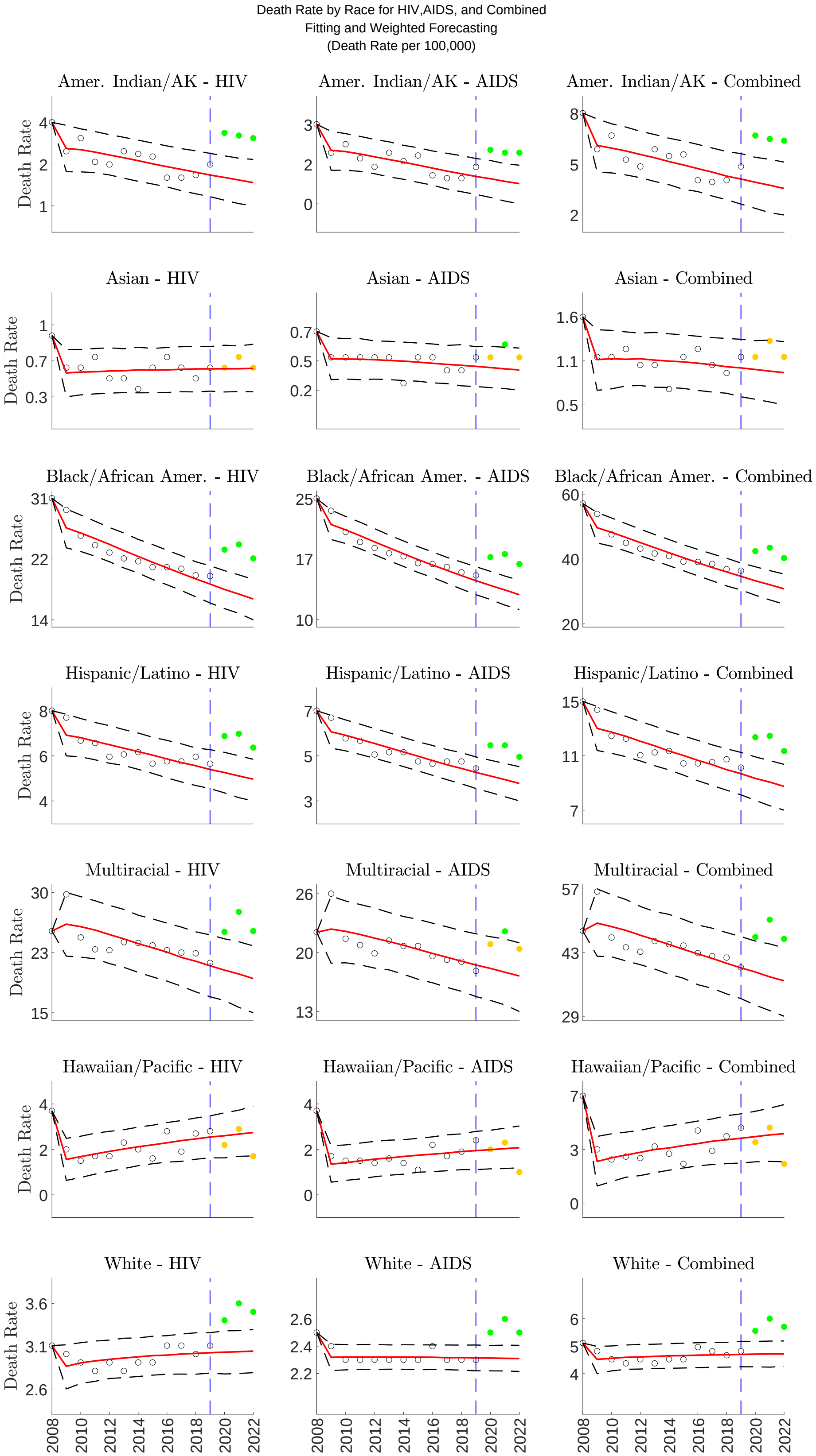


Figure S8: Death rate for HIV, AIDS, and HIV/AIDS combined by year using weighted ensemble model, stratified by race. This figure represents overall death rate in deaths per 100,000 people. The solid red line represents the modeled median death rate, with the dotted black line representing 95% Prediction Interval (PI). Data points shown in black represent observed data used for the calibration period, while solid-filled green/yellow data points in the years 2020-2022 represent forecasted data. Forecasted data in green represents excess death rates outside of the 95% PI, while forecasted data in yellow, if present, represents excess data rates within the 95% PI. The blue dotted vertical line denotes the year 2019, which is the end of the calibration period, and before COVID-19.

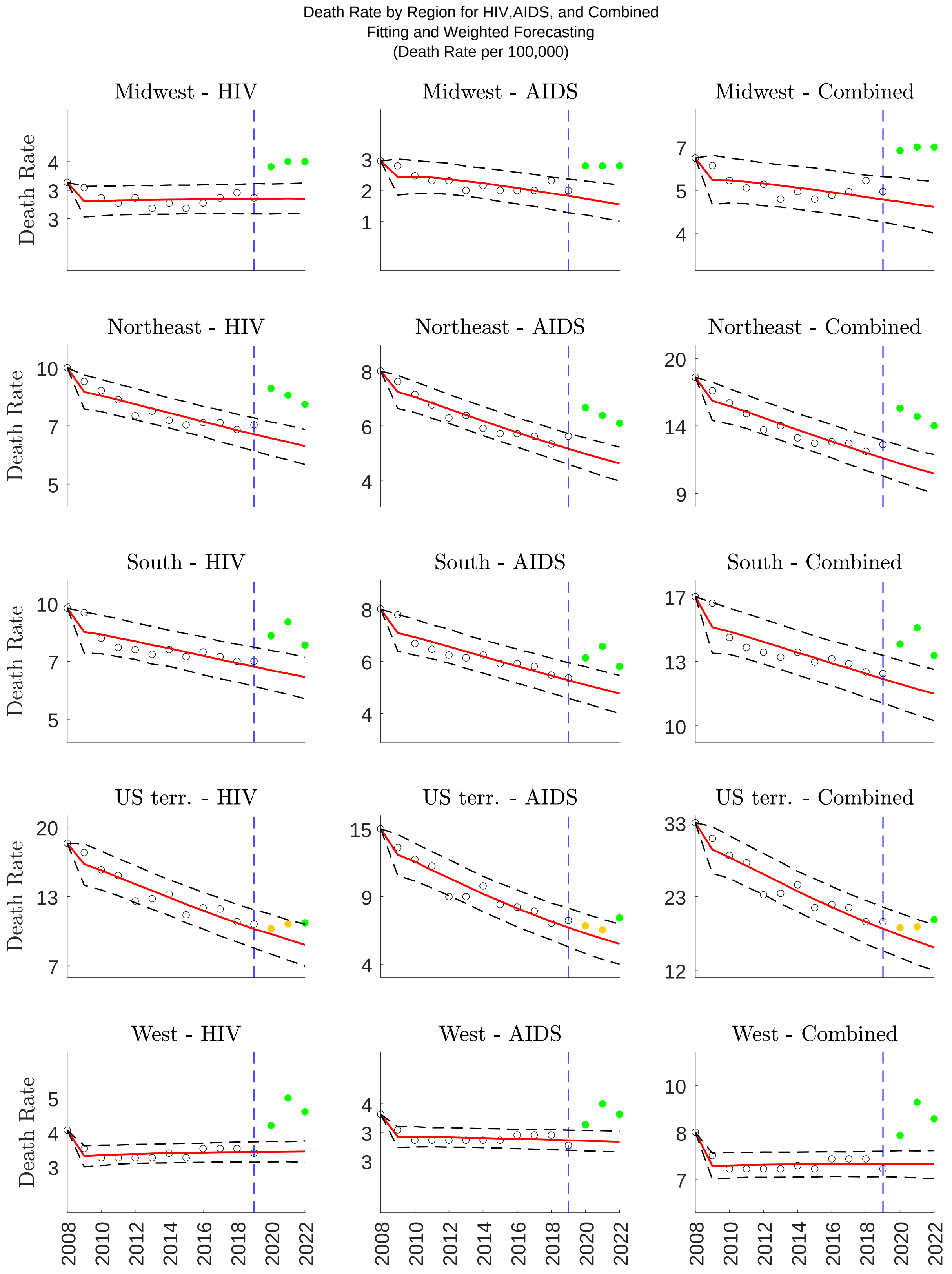


Figure S9: Death rate for HIV, AIDS, and HIV/AIDS combined by year using weighted ensemble model, stratified by region. This figure represents overall death rate in deaths per 100,000 people. The solid red line represents the modeled median death rate, with the dotted black line representing 95% Prediction Interval (PI). Data points shown in black represent observed data used for the calibration period, while solid-filled green/yellow data points in the years 2020-2022 represent forecasted data. Forecasted data in green represents excess death rates outside of the 95% PI, while forecasted data in yellow, if present, represents excess data rates within the 95% PI. The blue dotted vertical line denotes the year 2019, which is the end of the calibration period, and before COVID-19.

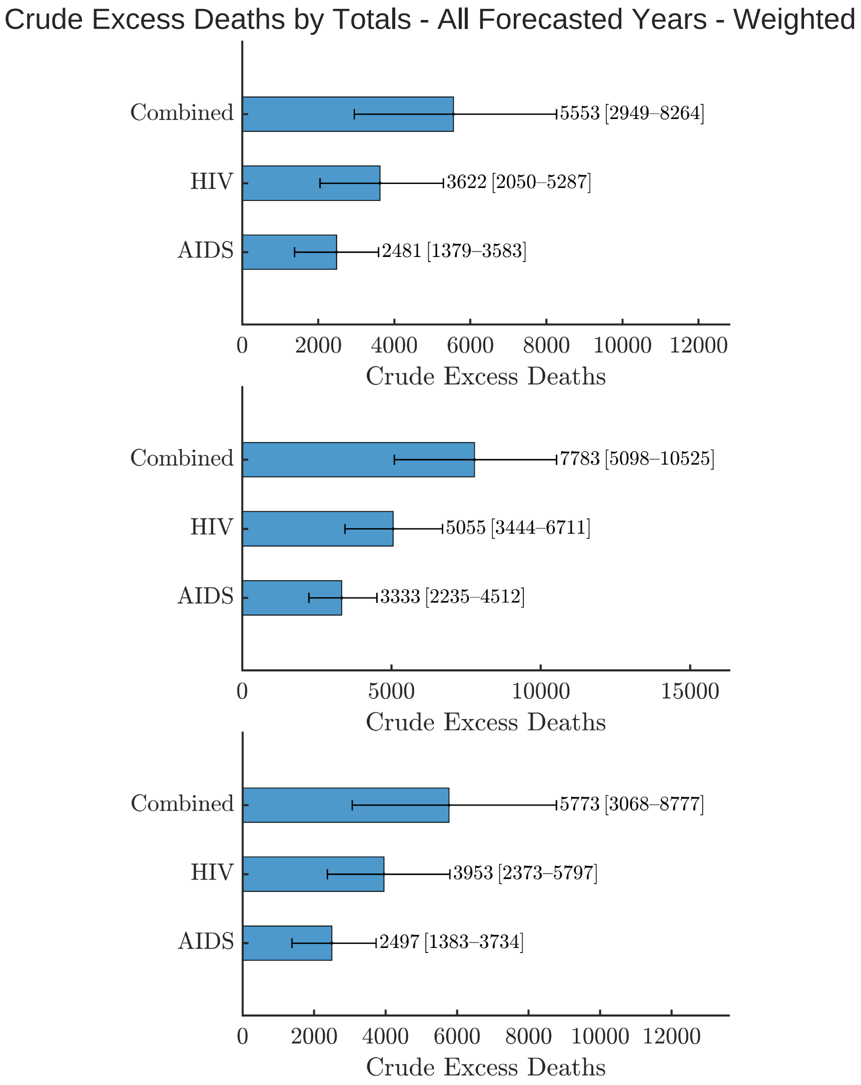


Figure S10: Crude excess deaths for HIV, AIDS, and HIV/AIDS combined 2020-2022 using weighted ensemble forecasting. This figure represents crude excess deaths. The 95% upper and lower bounds for median crude excess deaths are displayed in black bars.

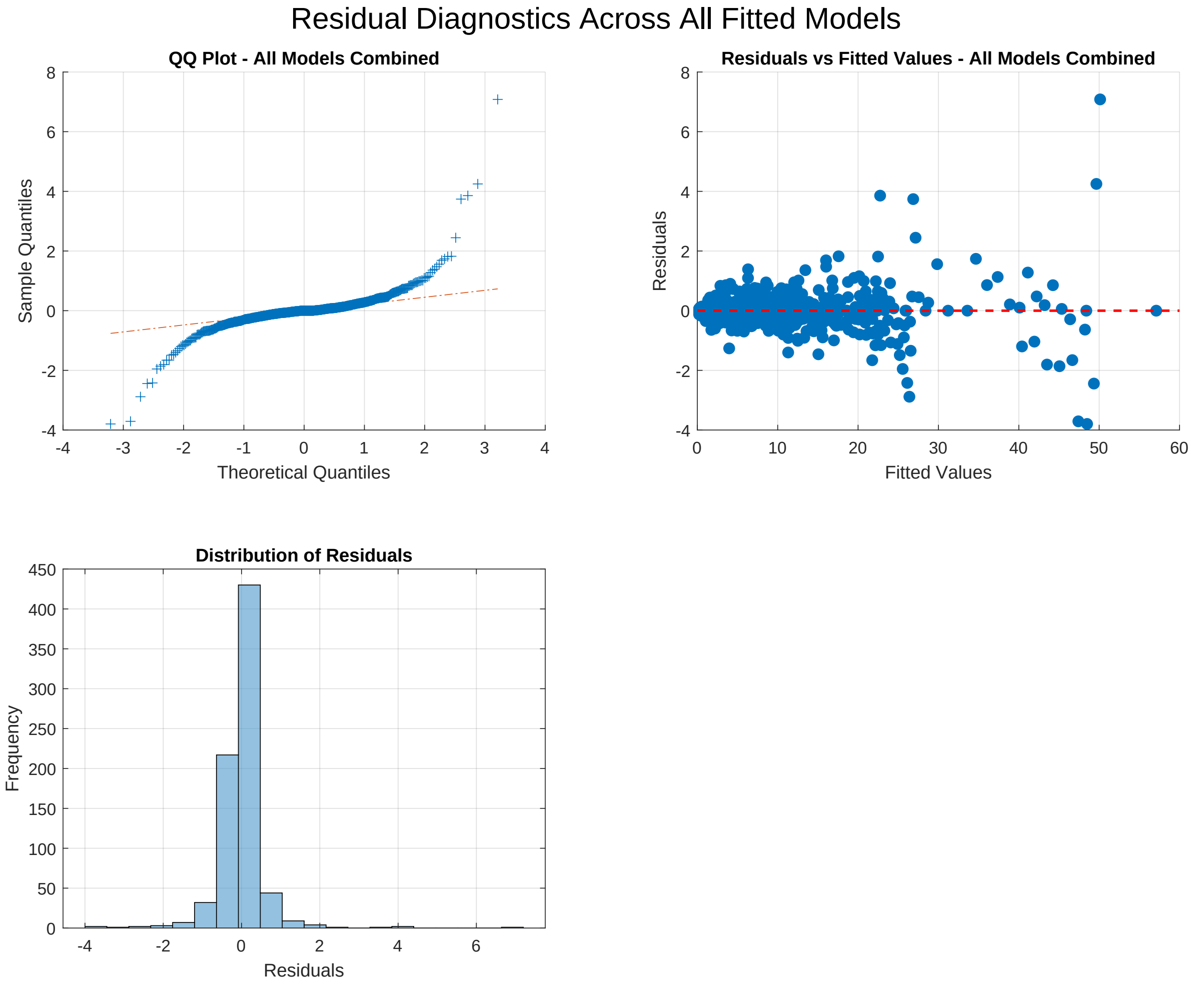


Figure S11: (A) Q–Q plot showing observed residuals versus theoretical quantiles from a normal distribution. (B) Residuals plotted against fitted values, with the red dashed line at zero indicating perfect prediction. (C) Histogram of residuals showing the distribution across all 756 observations (12 calibration years × 63 demographic-indicator combinations). Residuals approximate a normal distribution centered at zero with minor deviations at the extreme tails, supporting the validity of the assumed error structure.

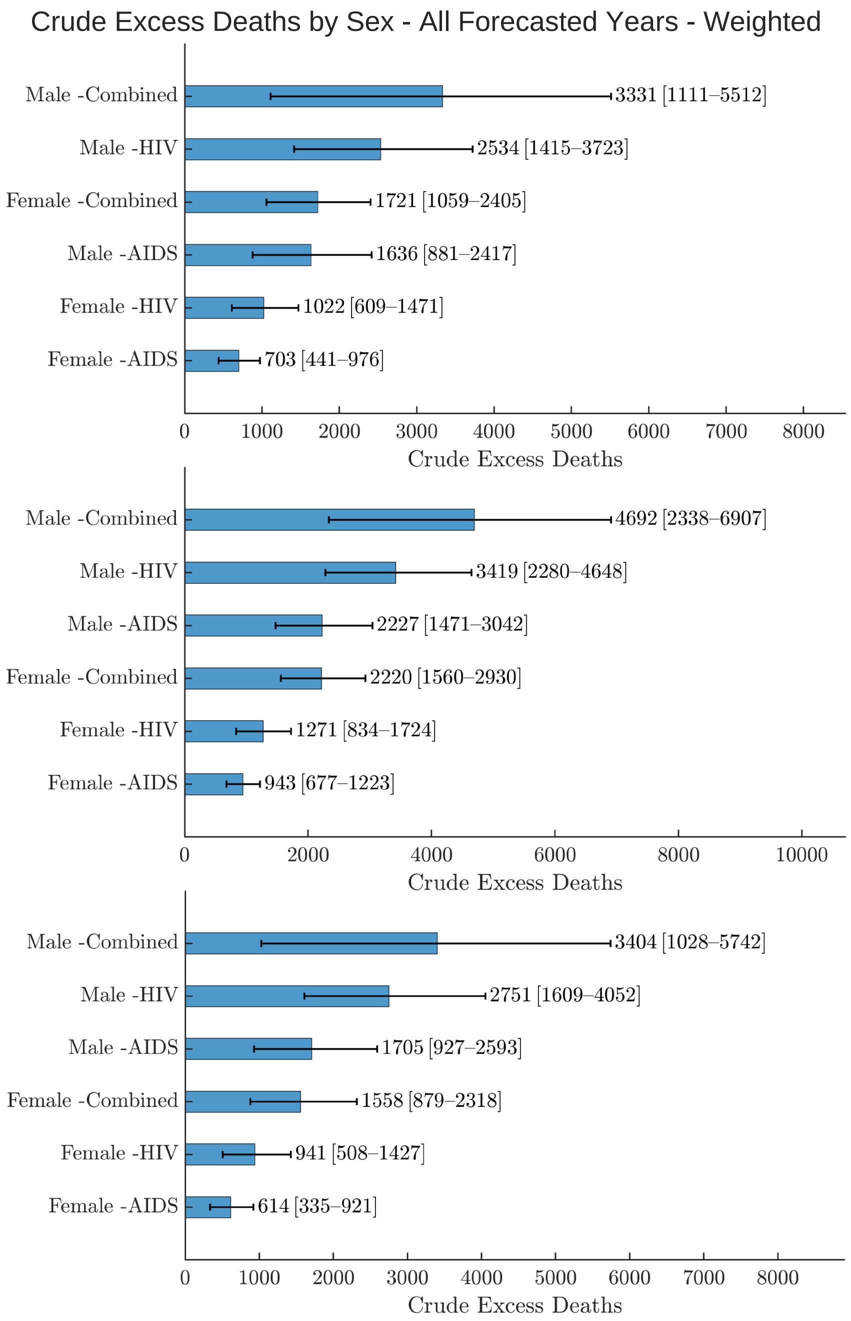


Figure S12: Crude excess deaths for HIV, AIDS, and HIV/AIDS combined for sex 2020-2022 using weighted ensemble forecasting. This figure represents crude excess deaths. The 95% upper and lower bounds for median crude excess deaths are displayed in black bars.

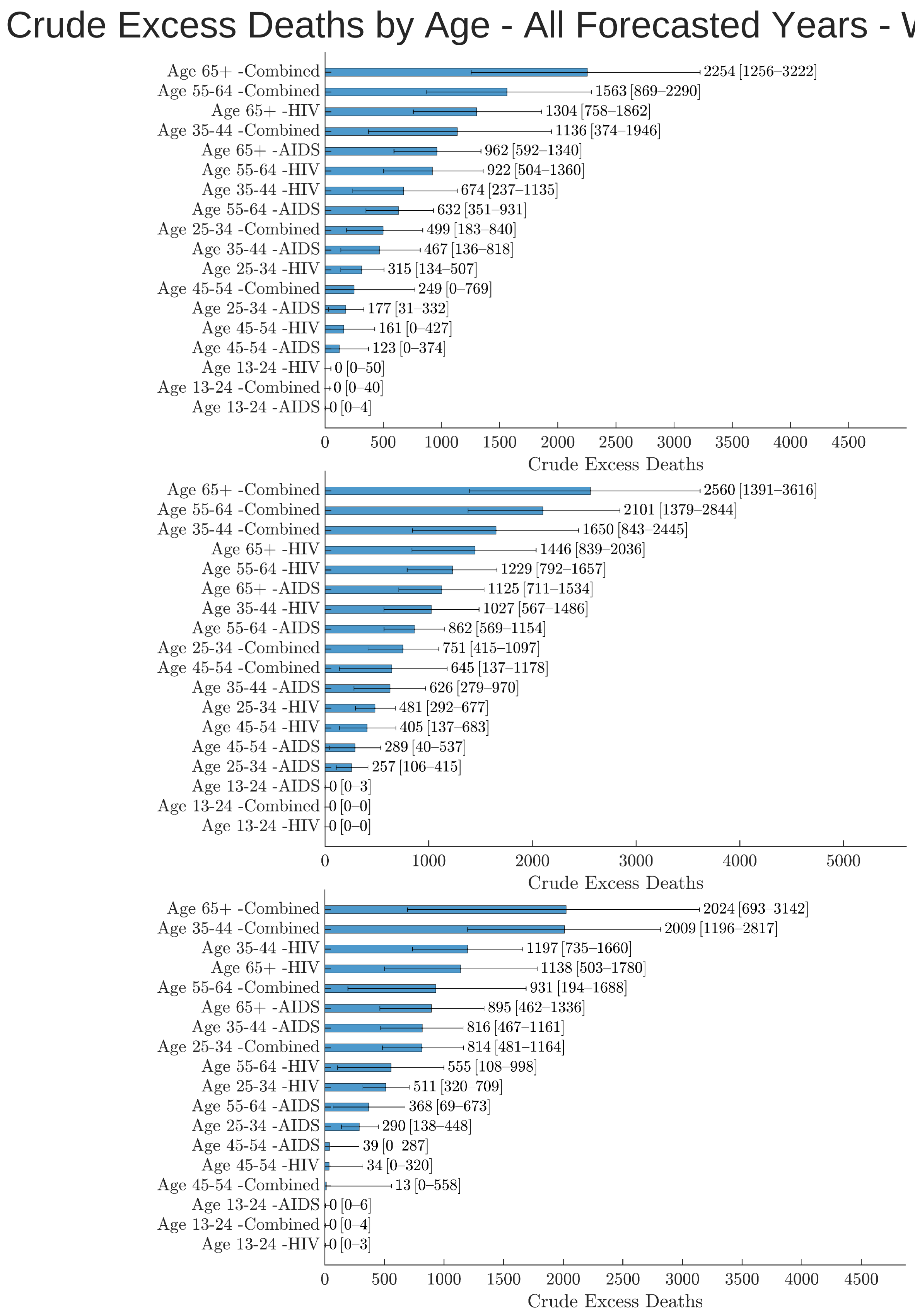


Figure S13: Crude excess deaths for HIV, AIDS, and HIV/AIDS combined for race 2020-2022 using weighted ensemble forecasting. This figure represents crude excess deaths. The 95% upper and lower bounds for median crude excess deaths are displayed in black bars.

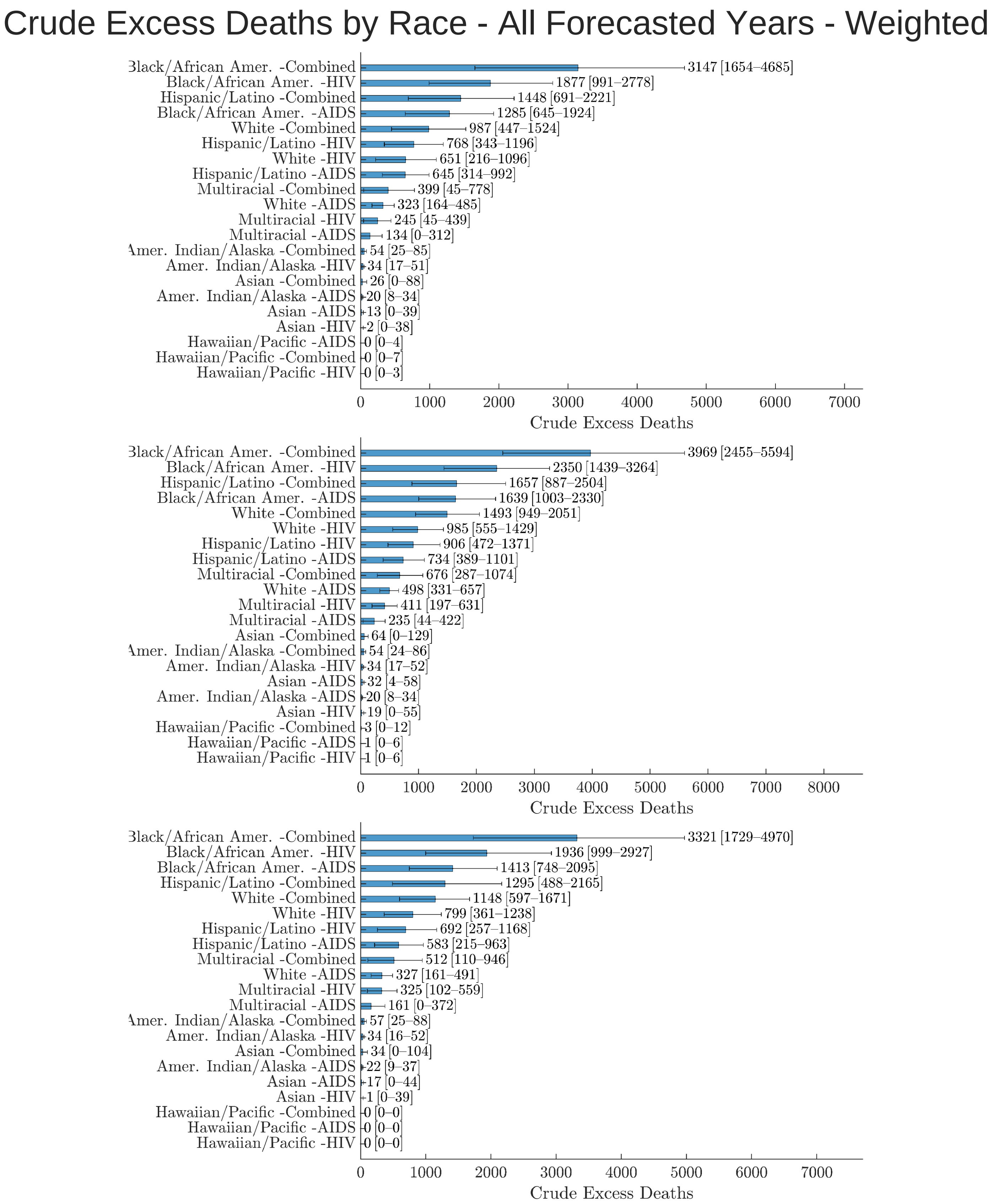


Figure S14: Crude excess deaths for HIV, AIDS, and HIV/AIDS combined for race 2020-2022 using weighted ensemble forecasting. This figure represents crude excess deaths. The 95% upper and lower bounds for median crude excess deaths are displayed in black bars.

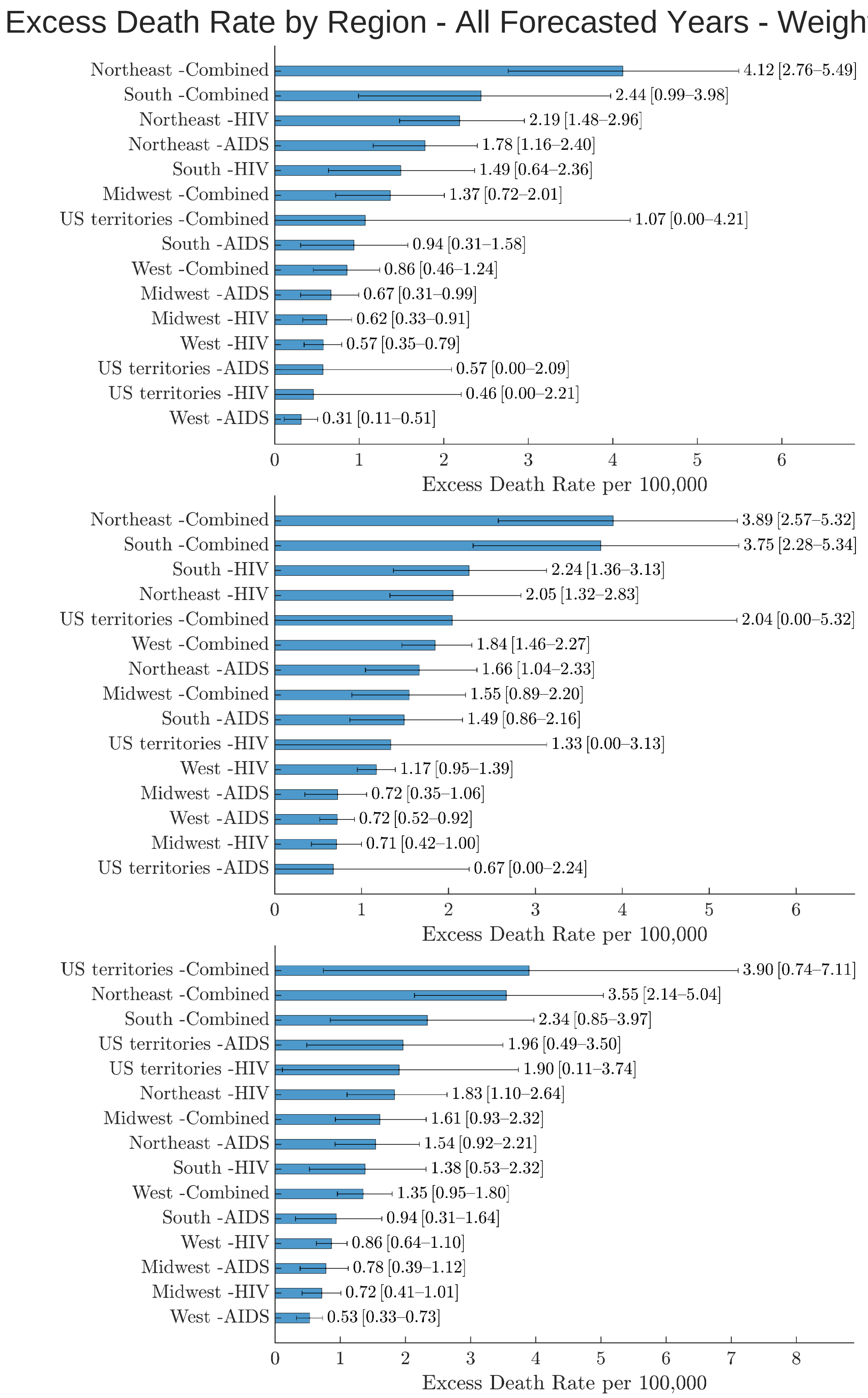


Figure S15: Excess deaths rate for HIV, AIDS, and HIV/AIDS combined for region 2020-2022 using weighted ensemble forecasting. This figure represents crude excess deaths. The 95% upper and lower bounds for median crude excess deaths are displayed in black bars.

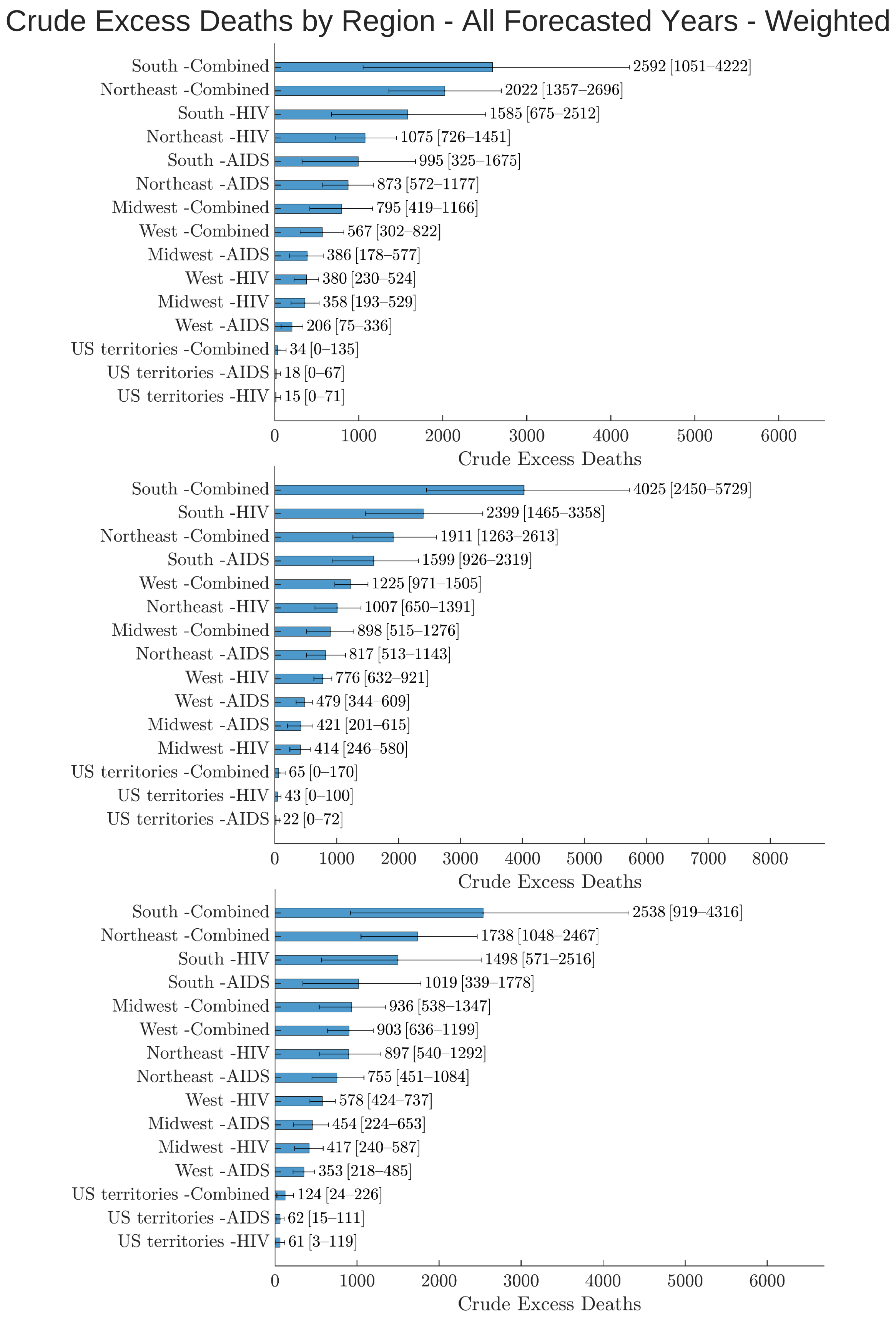


Figure S16: Crude Excess Deaths for HIV, AIDS, and HIV/AIDS combined for region 2020-2022 using weighted ensemble forecasting. This figure represents crude excess deaths. The 95% upper and lower bounds for median crude excess deaths are displayed in black bars.
